# Factors associated with home birth in Peru: An analysis of the Demographic and Health Survey, 2019

**DOI:** 10.1101/2021.06.01.21258107

**Authors:** Jackeline Huapaya-Torres, Yuly Santos-Rosales, Victor Moquillaza-Alcántara

## Abstract

**Objective:** To determine the proportion and factors associated with home birth in Peru, 2019.

**Material and methods:** Cross-sectional analytical design study where the 2019 Peruvian Demographic and Family Health Survey was analyzed. The association was evaluated using Poisson Regression, supplemented with the crude prevalence ratio (cPR) and adjusted (aPR).

**Results:** The records of 18,401 women were evaluated, where 5.39% (95%CI:4.83-6.03%) presented home birth. The probability of a home birth occurs increases when the pregnant woman is from the andean (aPR:1.24; 95%CI:1.02-1.48) and amazon region (aPR:1.38; 95%CI:1.16-1.64), resides in rural areas (aPR:3.34; 95%CI:2.61-4.29), presents less than 6 prenatal care (aPR:1.66; 95%CI:1.39 -1.96), it is very poor (aPR:9.62; 95%CI:5.13-18.1) or poor (aPR:2.39; 95%CI:1.26-4.52), it has not studied (aPR:2.66; 95%CI:2.02-3.50) or reached primary education (aPR:2.18: 95%CI:1.85-2.58) and has 2 children (aPR:1.64; 95%CI:1.46-1.85) or 3 or more children (aPR:2.18; 95%CI:1.67-2.87). On the other hand, having higher educational instruction (aPR:0.49; 95%CI:0.31-0.78) is associated with a lower probability of a home birth.

**Conclusions:** There is a low proportion of home births; however, this indicator increases significantly according to various geographical, sociodemographic, and obstetric factors that have been identified.

**HIGHLIGHTS:**

- The proportion of home births in Peru is low, although it reaches high values in the amazon and rural areas.
- Sociodemographic determinants such as poverty and low educational level are risk factors for home birth.
- Obstetric determinants such as having few prenatal controls or had having previously multiple deliveries are risk factors for home birth.

## INTRODUCTION

Delivery care must be humanized and have the assistance of a qualified professional, which can be done at home if the pregnancy is low risk. Currently, several countries have legislated home care, seeking to promote the autonomy of women during childbirth (1–3), thus avoiding medicalized, bureaucratic processes with little cultural adaptation, also favoring the satisfaction of the user when being cared for in the comfort of her home with greater control of this experience (4–7). Despite this, there are maternal and neonatal risks associated with home birth that should promote the formulation of new health strategies (7,8).

In countries with medium and low income, there have been efforts to favor institutional delivery; However, after a first hospital birth, women choose to practice home birth (9-11), which can be attributed to the poor cultural adaptation of the institutions, such as an act of protest or an appreciation of the guidance of women midwives and their partners (12). In Peru, even though the proportion of home births have been decreasing (13), the behavior is different inside the country, being more frequent in rural regions, which is characteristic of environments where there are remarkable economic gaps and social inequalities. (14,15).

The present study seeks to determine where the practice of home birth prevails, in order to focus support from the authorities. Also, it seeks to recognize the determinants that favor this practice, which will allow the generation of health strategies that strengthen the adaptation of institutional delivery to the own experiences of a community, which is not yet regulated in the country (16). Due to the aforementioned, the objective of the study was to determine the proportion and factors associated with home birth in Peru.

## MATERIALS AND METHODS

### Study desing

This report is a secondary base analysis of the Demographic and Family Health Survey (ENDES, for its acronym in Spanish) of Peru for the year 2019, with a cross-sectional analytical design.

### Data set

The database of the Demographic and Family Health Survey of 2019 was used, which is an annual survey carried out by the National Institute of Statistics and Informatics of Peru. The instrument is a public initiative that seeks to estimate indicators of the national budget programs, it also provides demographic information and, mainly health status of mother and children (17).

### Study sampling and participants

The survey selects its clusters (primary units) through a systematic random sampling, then the households (secondary units) are determined through balanced sampling. During 2019, the survey had 3 254 conglomerates, which contained 36 760 homes, distributed among department capitals, urban cities, and rural areas. These processes guarantee that it is the most representative survey at national level (17).

People that participated were women between the ages of 12 to 49 and children under 12 years old, who must stay overnight in the household, since the last night of being interviewed, giving a total of 40,809 participants. For this study, those who had not have a pregnancy during the last 5 years and presented incomplete data for all the questions considered as study variables were excluded, giving a sample of 18 401 complete records (**Figure 1**).

**Figure 1.**
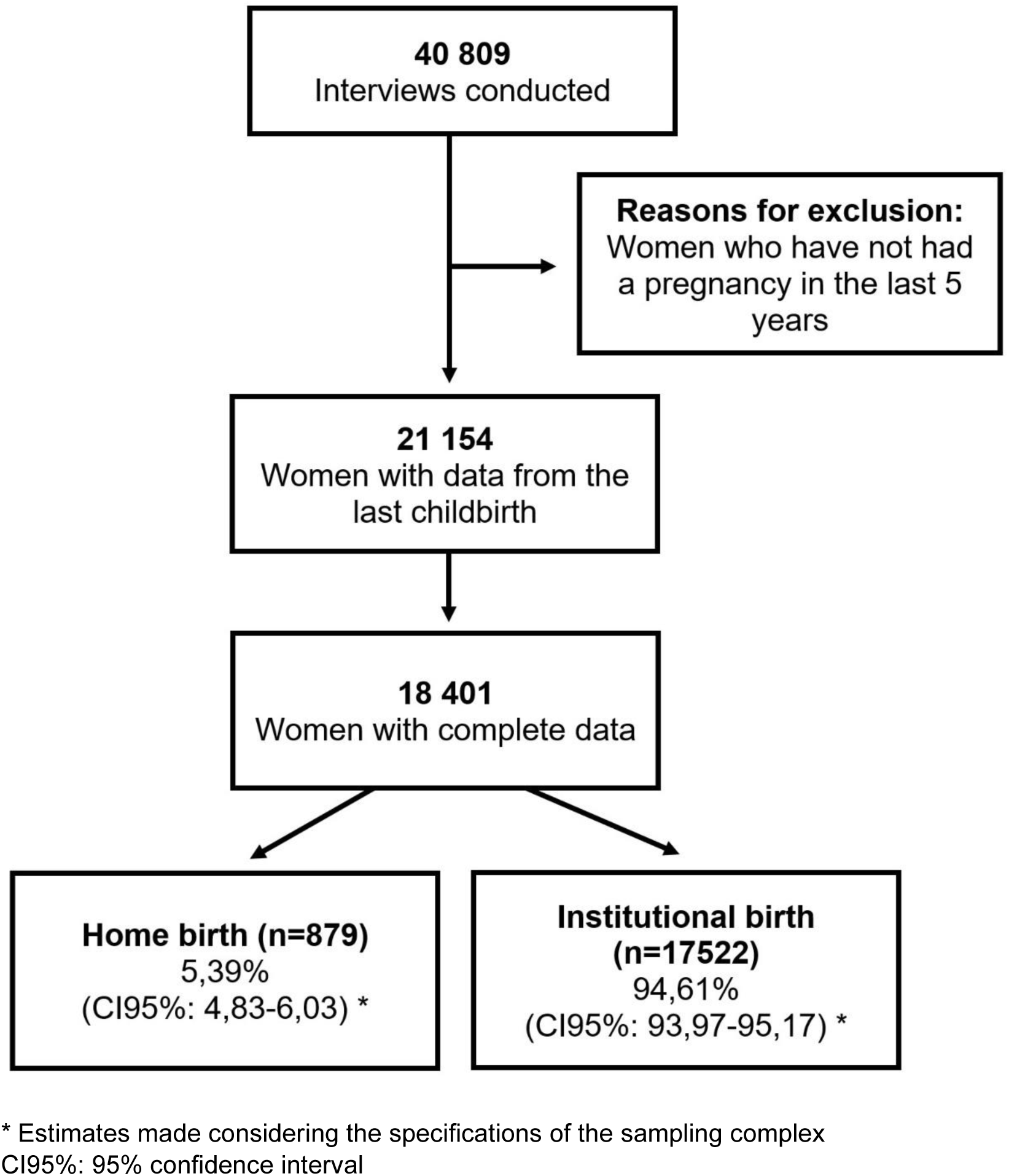
Participant selection flowchart.

### Measurements

The place where the last living child of the interviewed was born was considered as the dependent variable of the study. The question, assigned as 426A in the database, was “Where did she give birth to her (her last child)?” The responses were recategorized and it was coded as 1 when the participant indicated that her delivery was at home (own house or midwife’s house), in addition, it was coded as 0 when it was indicated that the delivery was institutional (health center, hospital, non-governmental organization or private establishment).

The independent variables considered for the study were the natural region where it comes from, the area of residence, the number of prenatal cares, trimester in which prenatal care begins, wealth index, highest educational level attained, age, birth order, newborn size, health insurance and presence of violence (emotional, physical, and sexual). The coding used in each variable of the study is found in **Table 1**.

**Table 1.** Description and analysis coding plan of selected variables of the study

|  | **Description and Categorization** | **Analysis Coding** |
| --- | --- | --- |
| **Outcome variable** |  |  |
| Home birth | Place where the interviewee gave birth to her last living child (11=Respondent's home, 12=Midwife's home, 21-27=Hospital, 31-33=Private institution, 41-42=Non-governmental organizations) | Options were recoded as follows:<br>11&12 as 1= Home birth<br>>12 as 0= Institutional birth |
| **Independent variables** |  |  |
| Natural region | Region where participant resides (1=Lima, 2=Rest of coast, 3=Andean, 4=Amazon) | Options were recoded as follows:<br>1&2 as 1= Coast<br>3 as 2= Andean<br>4 as 3= Amazon |
| Place of residence | Type of place of residence (1=Urban, 2=Rural) | Options were recoded as follows:<br>1 as 0= Urban<br>2 as 1= Rural |
| Antenatal visits | Total number of antenatal visits that the interviewee had during the pregnancy of her last living child (0=No antenatal visits, 1-97=same numeric value, 98=DK) | Options were recoded as follows:<br>0-5 as 1= <6<br>>6 as 0= 6 or more<br>98 was not considered for analysis |
| Timing of 1st antenatal check | Months of pregnancy that the interviewee was when she had her first antenatal visit (1-9=same numeric value, 98DK) | Options were recoded as follows:<br>1-3 as 0= First trimester<br>>3 as 1= >First trimester<br>98 was not considered for analysis |
| Wealth index | Wealth that the home has (1=Poorest, 2=Poorer, 3=Middle, 4=Richer, 5=Richest) | Used same coding |
| Highest educational level | Highest level of study passed (0=No education, 1=Primary, 2=Secondary, 3=Higher) | Used same coding |
| Age | Current age (0= Less than 1 year, 1-96=same numeric value, 97=>97, 98=DK) | Options were recoded as follows:<br><12 was not considered for analysis<br>12-17 as 1= 12-17 years<br>18-24 as 2= 18-24 years<br>25-34 as 3= 25-34 years<br>35-49 as 4= 35-49 years<br>>49 was not considered for analysis |
| Index to birth history | Serial number at birth of each child under 6 years of age (continuous) | Options were recoded as follows:<br>1= 1<br>2= 2<br>>2= 3 or more |
| Child size at birth | Size of child at birth (1=very large, 2=larger than average, 3=average, 4=smaller than average, 5=very small, 8 = DK) | Options were recoded as follows:<br>1&2 as 1= Large<br>3 as 2= Average<br>4&5 as 3= Small<br>8 was not considered for analysis |
| Affiliated with health insurance during pregnancy | Coverage of Health Insurance or Maternal-Infant Insurance during pregnancy (0=No, 1=Yes) | Used same coding |
| Emotional violence | Ever any emotional violence (0=No, 1=Yes) | Used same coding |
| Physical violence | Ever any physical violence (0=No, 1=Yes) | Used same coding |
| Sexual violence | Ever any sexual violence (0=No, 1=Yes) | Used same coding |

DK: Do not Know

### Statistical analysis

The analysis was made in STATA version 14 software, where the databases were downloaded in the beginning. Those are divided into chapters and the required ones for the study were joined by means of the “merge” command, considering as a binding variable the person identifier (assigned as “CASEID” in the database) (18). In the same way, all estimates considered the characteristics of the complex sampling presented by the survey.

Within the descriptive analysis, absolute frequencies and weighted proportions were determined according to complex sampling, accompanied by the 95% confidence interval. On the other hand, the characteristics of the participants were compared according to the place where their delivery occurred (bivariate analysis), the difference in the proportions was evaluated using Pearson’s Chi-square test, considering as associated that value of p less than 0.05.

Subsequently, a multivariate analysis was made through the Poisson Regression test, where the results were adjusted among all the variables that reached statistical significance. Crude Prevalence Ratios (cPR) and adjusted (aPR) were estimated with their respective 95% confidence interval and, finally, the p value was considered less than 0.05 like significant.

### Ethical review

Due to the study is a secondary base analysis with public access (http://iinei.inei.gob.pe/microdatos/), it does not merit the approval of an ethics committee.

## RESULTS

Among the 18 401 records analyzed, it is estimated that 5.39% (95%CI:4.83-6.03%) of women in Peru have a home birth, however, the proportion reaches 19.59% (95%CI:17.26-22.17%) in rural areas and 17.85% (95%CI:15.21-20.84%) in the Peruvian amazon. Within the sociodemographic characteristics, being very poor (21.91%), having no education (28.21%) or only primary education (20.64%) were the categories that presented high prevalence of home birth. On the other hand, when evaluating certain obstetric characteristics, it was found that women with 3 or more deliveries (21.40%) and those with a small newborn (7.20%) showed a higher proportion of home births. Regarding prenatal care, it was found that home birth is more frequent when pregnant women have less than 6 prenatal care (13.69%) and when they start after the first trimester of pregnancy (5.43%). (**Table 1**)

Next, a crude and adjusted analysis was carried out to evaluate which characteristics were associated with a home birth, which can be seen in **Table 2**. Risk factors that were found: living in the andean (aPR:1.24; p<0.001), amazon (aPR:1.38; p<0.001), or in rural areas (aPR:3.34; p<0.001), have less than 6 prenatal care (aPR:1.66: p<0.001), have a very poor wealth index (aPR:9.62; p<0.001) or poor (aPR:2.39; p=0.008), have no education (aPR:2.66; p<0.001) or have achieved primary education (aPR:2.18; p<0.001) and having had 2 (aPR:1.64; p<0.001) or more live births (aPR:2.18; p<0.001). On the other hand, having reached a higher education was shown to be a protective factor against home birth (aPR:0.49; p=0.003).

**Table 2.** Characteristics associated with presenting a home birth in the last delivery during the last 5 years in Peru, 2019

|  | Home birth |  |  |
| --- | --- | --- | --- |
|  | Absolute frequency<br>(n/N) | Weighted ratio *<br>(IC95%) | p † |
| **Natural region** |  |  |  |
| Coast | 204/8498 | 3.40 (2.67-4.32) | *Ref.* |
| Andean | 297/6233 | 6.92 (5.71-8.36) | <0.001 |
| Amazon | 378/3670 | 17.85 (15.21-20.84) | <0.001 |
| **Place of residence** |  |  |  |
| Urban | 133/13241 | 1.37 (1.08-1.72) | *Ref.* |
| Rural | 746/5160 | 19.59 (17.26-22.15) | <0.001 |
| **Antenatal visits** |  |  |  |
| < 6 | 247/1895 | 13.69 (11.52-14.55) | <0.001 |
| 6 or more | 632/16506 | 4.43 (3.94-4.98) | *Ref.* |
| **Timing of 1st antenatal check** |  |  |  |
| First trimester | 178/5305 | 3.74 (2.79-4.69) | *Ref.* |
| > First trimester | 618/13096 | 5.43 (5.06-5.80) | <0.001 |
| **Wealth index** |  |  |  |
| Poorest | 730/5095 | 21.91 (20.93-22.89) | <0.001 |
| Poorer | 109/5025 | 2.06 (1.82-2.30) | <0.001 |
| Middle | 31/3664 | 0.79 (0.53-1.05) | *Ref.* |
| Richer | 5/2709 | 0.21 (0.07-0.35) | 0.002 |
| Richest | 4/1908 | 0.19 (0.01-0.36) | 0.009 |
| **Highest educational level** |  |  |  |
| No education | 72/291 | 28.21 (23.55-32.87) | <0.001 |
| Primary | 454/3435 | 20.64 (19.45-21.83) | <0.001 |
| Secondary | 322/8577 | 4.09 (3.71-4.47) | *Ref.* |
| Higher | 31/6098 | 0.39 (0.23-0.56) | <0.001 |
| **Age** |  |  |  |
| 12 – 17 years | 25/270 | 10.12 (6.72-13.52) | *Ref.* |
| 18 – 24 years | 226/4193 | 7.37 (6.68-8.06) | 0.057 |
| 25 – 34 years | 368/8563 | 5.81 (5.38-6.24) | 0.002 |
| 35 – 49 years | 260/5375 | 6.22 (5.64-6.80) | 0.013 |
| **Index to birth history** |  |  |  |
| 1 | 765/16006 | 5.13 (5.10-5.16) | *Ref.* |
| 2 | 249/2270 | 11.27 (10.99-11.55) | <0.001 |
| 3 or more | 26/125 | 21.40 (19.33-23.47) | <0.001 |
| **Child size at birth** |  |  |  |
| Small | 259/3957 | 7.20 (6.42-7.98) | <0.001 |
| Average | 411/9546 | 4.97 (4.55-5.39) | *Ref.* |
| Large | 209/4898 | 4.76 (4.19-5.33) | 0.566 |
| **Affiliated with health insurance during pregnancy** |  |  |  |
| Yes | 781/13208 | 6.96 (6.56-7.36) | <0.001 |
| No | 98/5193 | 2.05 (1.68-2.42) | *Ref.* |
| **Emotional violence** |  |  |  |
| Yes | 169/3980 | 4.73 (4.04-5.42) | 0.002 |
| No | 742/14421 | 5.94 (5.55-6.33) | *Ref.* |
| **Physical violence** |  |  |  |
| Yes | 226/4870 | 5.31 (4.66-5.96) | 0.069 |
| No | 685/13531 | 5.81 (5.40-6.22) | *Ref.* |
| **Sexual violence** |  |  |  |
| Yes | 59/843 | 7.42 (5.64-9.20) | 0.118 |
| No | 852/17499 | 5.59 (0.49-6.31) | *Ref.* |

95%CI: 95% confidence interval; *Ref.*: Reference category
\* The weightings were carried out considering the characteristics of the complex sampling
† Evaluated with Poisson regression with sampling weighting.

**Table 3.** Characteristics associated with presenting a home birth in the last delivery during the last 5 years in Peru, 2019

|  | Home birth |  |  |  |
| --- | --- | --- | --- | --- |
|  | cPR (CI95%) | p † | aPR (CI95%) | p † |
| **Natural region** |  |  |  |  |
| Coast | Ref. |  | Ref. |  |
| Andean | 2.11 (1.53-2.91) | <0.001 | 1,24 (1,02-1,48) | <0,001 |
| Amazon | 6.17 (4.51-8.45) | <0.001 | 1,38 (1,16-1,64) | <0,001 |
| **Place of residence** |  |  |  |  |
| Urban | Ref. |  | Ref. |  |
| Rural | 17.6 (13.3-23.3) | <0.001 | 3,34 (2,61-4,29) | <0,001 |
| **Antenatal visits** |  |  |  |  |
| < 6 | 3.42 (2.80-4.17) | <0.001 | 1,66 (1,39-1,96) | <0,001 |
| 6 or more | Ref. |  | Ref. |  |
| **Timing of 1st antenatal check** |  |  |  |  |
| First trimester | Ref. |  | Ref. |  |
| > First trimester | 1.47 (1.19-1.82) | <0.001 | 0,99 (0,84-1,16) | 0,881 |
| **Wealth index** |  |  |  |  |
| Poorest | 24.3 (17.0-34.6) | <0.001 | 9,62 (5,13-18,1) | <0,001 |
| Poorer | 2.56 (1.73-3.81) | <0.001 | 2,39 (1,26-4,52) | 0,008 |
| Middle | Ref. |  | Ref. |  |
| Richer | 0.22 (0.08-0.56) | 0.002 | 0,32 (0,07-1,48) | 0,147 |
| Richest | 0.25 (0.09-0.70) | 0.009 | 0,86 (0,23-3,27) | 0,824 |
| **Highest educational level** |  |  |  |  |
| No education | 6.60 (5.35-8.16) | <0.001 | 2,66 (2,02-3,50) | <0,001 |
| Primary | 4.75 (4.21-5.36) | <0.001 | 2,18 (1,85-2,58) | <0,001 |
| Secondary | Ref. |  | Ref. |  |
| Higher | 0.13 (0.09-0.19) | <0.001 | 0,49 (0,31-0,78) | 0,003 |
| **Age** |  |  |  |  |
| 12 – 17 years | Ref. |  | Ref. |  |
| 18 – 24 years | 0.69 (0.47-1.01) | 0.057 | 1,18 (0,67-2,10) | 0,568 |
| 25 – 34 years | 0.56 (0.38-0.82) | 0.002 | 1,16 (0,66-2,05) | 0,613 |
| 35 – 49 years | 0.62 (0.42-0.90) | 0.013 | 1,05 (0,59-1,86) | 0,877 |
| **Index to birth history** |  |  |  |  |
| 1 | *Ref.* |  | *Ref.* |  |
| 2 | 2.29 (2.02-2.60) | <0.001 | 1,64 (1,46-1,85) | <0,001 |
| 3 or more | 4.39 (3.17-6.08) | <0.001 | 2,18 (1,67-2,87) | <0,001 |
| **Child size at birth** |  |  |  |  |
| Small | 1.59 (1.40-1.81) | <0.001 | 1,03 (0,88-1,20) | 0,710 |
| Average | *Ref.* |  | *Ref.* |  |
| Large | 1.04 (0.91-1.19) | 0.566 | 0,95 (0,80-1,12) | 0,541 |
| **Affiliated with health insurance during pregnancy** |  |  |  |  |
| Yes | 3.13 (2.55-3.86) | <0.001 | 0,85 (0,67-1,10) | 0,215 |
| No | *Ref.* |  | *Ref.* |  |
| **Emotional violence** |  |  |  |  |
| Yes | 0.78 (0.67-0.92) | 0.002 | 0,93 (0,75-1,15) | 0,516 |
| No | *Ref.* |  | *Ref.* |  |
| **Physical violence** |  |  |  |  |
| Yes | 0.88 (0.76-1.00) | 0.069 | 0,93 (0,76-1,12) | 0,440 |
| No | *Ref.* |  | *Ref.* |  |
| **Sexual violence** |  |  |  |  |
| Yes | 1.22 (0.95-1.57) | 0.118 | 1,15 (0,84-1,58) | 0,389 |
| No | *Ref.* |  | *Ref.* |  |

† Evaluated with Poisson regression with sampling weighting.
cPR y aPR: Crude and adjusted prevalence ratio, 95%IC: 95% confidence interval; *Ref.*: Reference category.

## DISCUSSION

This study sought to determine those factors that are associated with the practice of home birth in Peru, owing to, despite the fact that the proportion has been decreasing, this reduction is not homogeneous in all regions of the country. It was found that those regions that are part of the Peruvian amazon and Andes are more likely to present cases of home births, which matches with a similar study that had the same geographical area (19). It should be noted that these regions are characterized by sociodemographic hurdle, which leads to the inaccessibility of health services (20–22). According to many authors, other reasons why women in the Peruvian Amazon prefer to have a home birth is the cultural pattern and the lack of cohesiveness with health facilities (23), which is why it is suggested to continue promoting the adequacy of intercultural delivery.

On the other hand, the present study showed that women living in rural areas are more likely to have home births, which is consistent with a recent systematic review (24). One possible explanation lies in the fact that pregnant women in rural areas feel fear that their privacy and traditions are not going to be respected in health facilities, and some indicated that they were afraid of being victims of abuse by health professionals (25–27), which highlights the marked gap between the rural population and health care.

Having less than 6 prenatal care (APN) makes it difficult to monitor the pregnant woman, preventing screening for lot of complications (28–30) and raising awareness of the importance of an institutional delivery (31). It should be noted that the number of APN alone does not determine the type of delivery, because the decision is multifactorial (32,33). However, the presence of a non-refocused routine APN, which is one where the birth plan, family education and multisectoral work with community leaders are not given importance (34), favors migration to home birth. On the other hand, the evidence shows that the absence of prenatal care or its beginning after 16 weeks widely increases the probability of a home birth, which can be explained by the limited information and procedures that can be developed over time rest that remain, reducing the expected quality in APNs (35).

Considering the wealth index, being very poor or poor increases the probability of having a home birth, which is similar to studies carried out in Tanzania (36) and Ethiopia (37). This might be due to the difficulties inherent to limited income, which creates a gap to pay for transportation, medicines, and medical care (38). Given this, it is the government that must guarantee that the necessary means are generated for the development of prenatal, delivery and postpartum care for those women who do not have the necessary resources (39).

It was found that having no education or having primary education increases the probability of a home birth, while having a higher education level reduces this probability. Data that is similar to those found in Peruvian studies (19) and in African countries, where reinforces the need to implement educational programs, that emphasizes the importance of institutional care and friendly services focused on those with limited economic resources, during pregnancy. (36,40). On the other hand, women with higher education usually have access to information about the risks or complications for the mother and her baby and value the importance of the APN, as well as greater purchasing power that allows them to pay for the services described in the previous paragraph (41–43).

Finally, the study determined that as there are more living children, the probability of having a home birth increases. This premise has also been determined in previous studies, which is explained by the bad experience that pregnant women have inside the institutions, which motivates them to change the place of delivery (44). In this regard, initiatives have been proposed that incorporate traditional midwives within (inside) the institution, where they can establish educational discussions, due to it has been identified that the usual clinical environment does not allow to establish appropriate communication (45–47). These proposals can be applied to improve care, which would improve adherence to institutional care, where there are tools to sort out complications in a timely manner.

The results of the study need to be limited to certain limitations. In the first place, there is the possibility that the interviewees do not remember certain characteristics of an event that could have happened during the last 5 years. Also, the cross-sectional analysis of the variables does not allow to establish a direct causality, but only a link that needs to be explored in future studies.

It is concluded that there are determinants that favors the occurrence of home births. It is necessary to approach a promotional preventive work from educational institutions and within families, so with that a pregnancy would be planned and its prenatal care will begin in a timely manner before 13 weeks, likewise, that the personal who provides the care are qualified and offer a refocused prenatal care, considering interculturality, multisectorality and that work is coordinated with the different social agents.

## Data Availability

The data is publicly accessible and can be obtained at the following link: http://iinei.inei.gob.pe/microdatos/

## ACKNOWLEDGEMENTS

To Claudia Ordoñez-Vargas and Kervyn Ynocente-Lazares for reviewing and correcting the translation of the article.

## Notes

**Financing:** This research did not receive any specific grant from funding agencies in the public, commercial, or not-for-profit sectors.

### Competing Interest Statement

The authors have declared no competing interest.

### Funding Statement

This research did not receive any specific grant from funding agencies in the public, commercial, or not-for-profit sectors

### Author Declarations

Due to the study is a secondary base analysis with public access, it does not merit the approval of an ethics committee.

